# Reconstructing pathogen-specific antibody binding epitopes and age-dependent immune signatures from proteomic-scale peptide libraries

**DOI:** 10.1101/2025.09.21.25336286

**Authors:** Everlyn Kamau, Nikolina Walas, Minlu Zhang, Kathy Kamath, Jack Reifert, John Shon, Shahjahan Ali, Md Ziaur Rahman, Abul K. Shoab, Syeda L. Famida, Salma Akther, Md. Saheen Hossen, Palash Mutsuddi, Mahbubur Rahman, Andrew N. Mertens, Richelle C. Charles, Daniel T. Leung, Stephen Luby, Audrie Lin, Benjamin F. Arnold

**Affiliations:** Francis I. Proctor Foundation, University of California, San Francisco, California, USA; School of Public Health, University of California, Berkeley, California, USA; Serimmune Inc., California, USA; Department of Epidemiology, Colorado School of Public Health, Aurora, Colorado, USA; Environmental Health and WASH, Health System and Population Studies Division, International Centre for Diarrhoeal Disease Research, Dhaka, Bangladesh; Global Health and Migration Unit, Department of Women’s and Children’s Health, Uppsala University, Sweden; Division of Epidemiology, University of California, Berkeley, California, USA; Massachusetts General Hospital, Harvard Medical School, Harvard T.H. Chan School of Public Health, Boston, USA; Division of Infectious Diseases, Department of Internal Medicine, University of Utah, Salt Lake City, USA; Division of Infectious Diseases and Geographic Medicine, Stanford University, Stanford, USA; Department of Microbiology and Environmental Toxicology, University of California, Santa Cruz, California, USA; Department of Ophthalmology, University of California, San Francisco, California, USA; Institute for Global Health Sciences, University of California, San Francisco, California, USA

## Abstract

Public health interventions involving improved water, sanitation and promotion of hygiene behaviors (WASH) plus nutrition, were implemented in Bangladesh in a large randomized controlled trial to assess impact on childhood enteric infection and diarrheal disease. Here, we evaluated magnitude and breadth of humoral responses to enteric pathogens in a subset of children among those received intervention (n=60) versus a control group (n=60) using an integrated method of bacterial display peptide library screening, next-generation sequencing and computational analysis to characterize individual antibody repertoires in serum collected at median ages of 3, 14 and 28 months. We determined high seroprevalence for enteric infections and show that antibody recognition of the putative epitopes and antigenic regions remained consistent over time. With mathematical models, we inferred waning of maternal immunity, and a subsequent immunity boost due to infection. Random peptide library screening has potential utility for rigorous analysis of antibody responses and identification of epitopes indicative of protective humoral immune responses.

**Summary:** Unbiased random peptide assay provided high resolution data on immune signatures and antibody dynamics for enteric infections in infancy and childhood.

## INTRODUCTION

Diarrheal disease accounts for substantial childhood morbidity and mortality with more than half a million estimated deaths and more than four episodes per child per year (Global Burden of Diseases, Injuries, and Risk Factors Study (GBD) 2017, ^1–3^). Viral, bacterial, and parasitic enteropathogens, including enterotoxigenic *Escherichia coli (E. coli)*, Rotavirus, *Shigella* spp, *Campylobacter* spp, Norovirus, astrovirus, and *Cryptosporidium* spp, are leading causes of diarrheal illness and diarrheal-associated deaths in children living in low- and middle-income countries. In these settings, enteropathogen exposure is intense with the primary window of infection at less than three years of age ^4–6^.

When used to study population-level transmission, serology or antibody-based measurements provide useful endpoints in intervention studies or randomized control trial designs with infrequent sampling or without continuous surveillance ^7^. This is because the immunologic signal detects infections that begin and resolve between sampling points. Antibody levels integrate exposure over time and embed information about infections in the intervals between measurements, therefore complementing pathogen detection to provide a more complete history of infection.

Phage and bacterial display assays present large libraries of potential peptide epitopes in tiled arrays to facilitate comprehensive mapping of all potential antibody recognition sites. This offers an advantage over traditional and other high throughput serological techniques such as ELISA, lateral flow assays and multiplex bead assays, which cannot readily or systematically achieve similar level of mapping ^8,9^. These highly multiplexed assays leverage high-throughput sequencing to profile antibody repertoires against entire pathogen proteomes without purifying the target proteins or analyzing each peptide individually ^8,10^, and individual- and group-specific serological profiles can be defined comparatively between those with and without disease or with and without infection ^11–13^.

Here, we used a random bacterial display peptide library to determine serological profiles of various target enteric pathogens in the context of a household water, sanitation, hygiene (WASH) and nutritional intervention. The random peptide library allowed for tiling against arbitrary proteomes, whereas most phage display libraries would typically require selection of organisms prior to library construction. The WASH Benefits cluster randomized trial in rural Bangladesh was conducted to assess, among other questions, whether improved WASH delivered to household compounds reduced diarrhea in a birth cohort compared to standard practices ^14^. The trial enrolled 5,551 pregnant women and evaluated outcomes at 1-year and 2-years’ follow-up including caregiver-reported diarrhea in the past 7 days among children who were in utero or younger than 3 years at enrolment ^14^. The trial reported a 7-day diarrhea prevalence of 3.5% among index children and children under 3 years at enrolment who received combined WASH and nutrition (WASH+N), lower compared to a prevalence of 5·7% in the control group. Nutritional strategies including supplementation have been suggested to increase resistance to infections and are recommended for improving public health ^1516^.

We therefore hypothesized that (i) the WASH+N intervention would decrease enteric infections leading to lower enteric pathogen immune responses compared to the control group; (ii) and that if children in the intervention group experienced fewer infections, then they could exhibit narrower breadth of immunoglobulin G (IgG) response at the epitope level compared with the control group who may have been infected multiple times by the same pathogen. Conversely, if repeated infections led to a higher specificity of antibody response, then the control group may exhibit a narrower breadth (fewer IgG epitope hits).

## METHODS

### Ethics statement

Participants provided written, informed consent before enrollment and before specimen collection. The protocol of the original study (Clinical Trial Registration NCT01590095) was approved by Ethical Review Committees at the International Centre for Diarrheal Disease Research, Bangladesh (PR-11063), the Committee for the Protection of Human Subjects at University of California, Berkeley (2011-09-3652), the Institutional Review Board at Stanford University (25863) and at the University of California, San Francisco (22-36722).

### Study design and eligibility

The WASH Benefits Bangladesh cluster-randomized trial was designed to generate evidence about the impact of sanitation, water quality, handwashing, and nutrition interventions on growth and development in early childhood ^17^. The trial had seven arms - chlorinated drinking water (water); upgraded sanitation (sanitation); promotion of handwashing with soap (handwashing); combined water, sanitation, and handwashing; counselling on appropriate child nutrition plus lipid-based nutrient supplements (nutrition); combined water, sanitation, handwashing, and nutrition; and control (data collection only). The primary trial outcomes were caregiver-reported diarrhea in the past 7 days among children who were in utero or younger than 3 years at enrolment and linear growth (length-for-age Z score among children born to enrolled pregnant women) ^14^. Here, we analyzed venous blood samples collected longitudinally in the trial at the median ages of 3, 14, and 28 months in a substudy from the control and combined WASH+N (“intervention”) groups to measure the effect of the combined intervention on immune responses among children ^18^. Sample collection occurred in 2013 and 2014. Our analysis included children in the two groups with blood samples collected at three follow-up visits – “visit 1”: median age 3 months, “visit 2”: median age 14 months, and “visit 3”: median age 28 months.

We applied two restrictions to improve balance between the two arms with respect to measurement dates and age at each visit. First, we excluded children whose first measurement was taken between August and November 2013 to improve comparability between arms in sampling timing with respect to calendar date and season, since measurement in control clusters was paused during that period due to political instability in Bangladesh. Second, we limited the cohort to children who were between 28 and 200 days old at visit 1, and between 365 and 548 days (1 to 1.5 years old) at visit 2. Conservatively assuming 50% seroprevalence in the control group at age 14 months, we determined that a sample size of 60 children per group would have at least 80% power to detect an absolute reduction of 25% seroprevalence (from 50% to 25%) with a two-sided *α* = 0.05 using a standard equation for comparison of two proportions. Power for the expected quantitative outcomes could be higher but due to the novelty of the outcome measurement, there were no available data to inform power calculations. Therefore, among 147 children who met the inclusion and exclusion criteria, we selected a random sample of 120 children (60 per group, 360 total samples) for analysis (**Fig S1**). Child age and sex were well balanced between groups, with slightly more female children and slightly older measurement ages at visits 1 and 2 in the control group (**Table S1)**.

It should be noted that at visit 2, several enteropathogens (bacteria, viruses, and parasites) were previously measured in fecal samples via quantitative polymerase chain reaction (PCR) using a TaqMan array card ^19^. Their prevalence among the 120 children enrolled in this substudy is shown in **Table S2**.

### Serum antibody profiling using bacterial display libraries

#### I. Library preparation and selection

Detection of the antibody repertoire composition was done using the serum antibody repertoire analysis (SERA) method published previously ^20,21^ (**Fig 1)**. Briefly, *E. coli* expressing random 12-mer peptide display libraries were induced for peptide expression, resuspended in 15% glycerol/PBS (PBSG) and aliquoted in 96-well deep-well plates at 10-fold the library diversity in each well (8×10^10^ cells per well). Library selection was performed by incubating serum to a final 1:25 dilution and the antibody-bound bacterial clones captured using Protein A/G Sera-Mag SpeedBeads (GE Life Sciences, 17152104010350) (6.25% the beads’ stock concentration). The selected bacterial pools were resuspended in growth medium and incubated overnight at 37 °C with shaking at 300 rpm for clone propagation.

**Fig 1.**
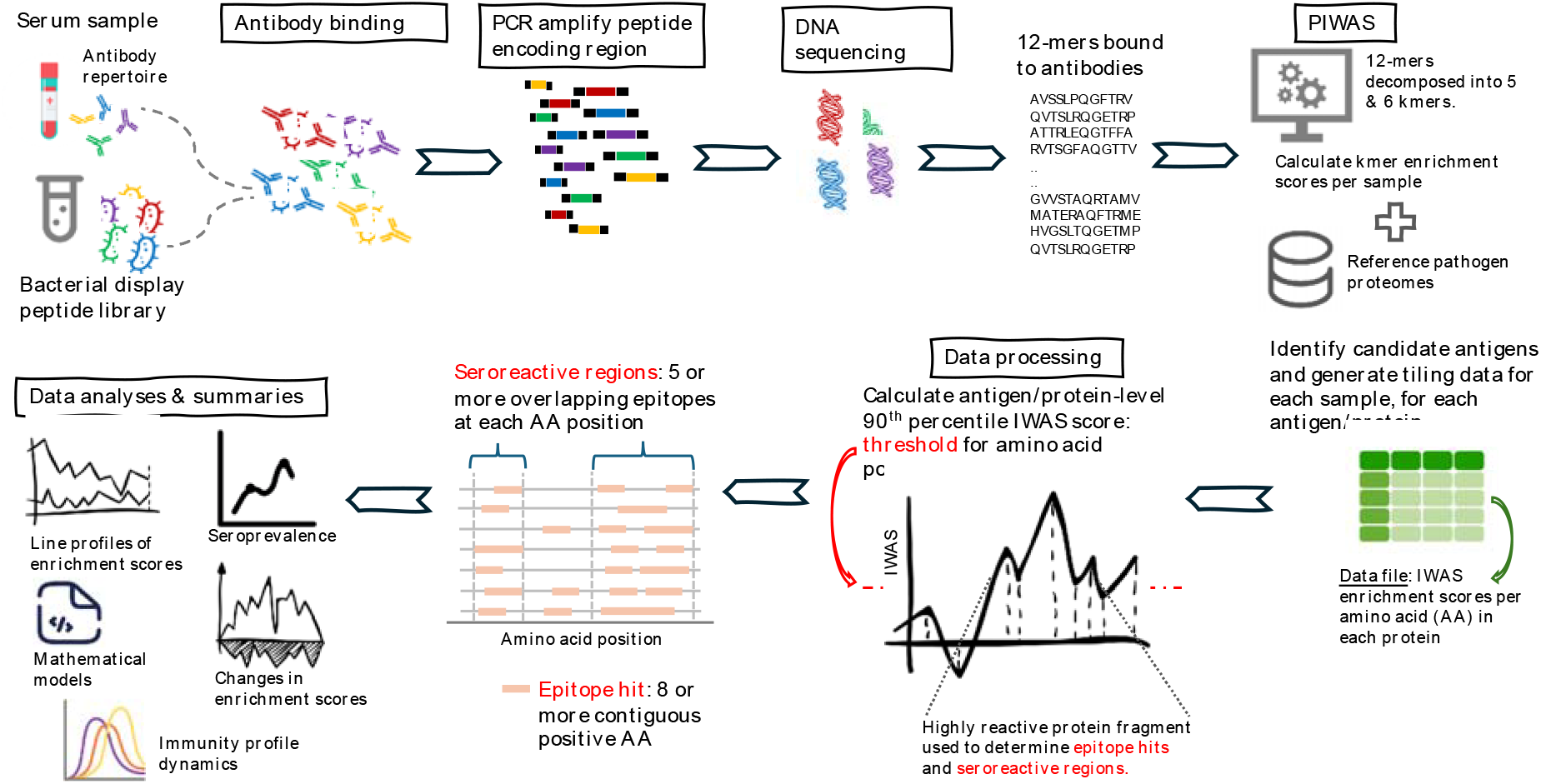
Sample processing and bioinformatic analysis. In the serum epitope repertoire analysis (SERA), serum samples were mixed with a fully random 12-mer peptide library expressed on the surface of *E. coli* bacteria, followed by isolation of the bacteria-antibody complexes using magnetic beads. Sequencing data from each sample were obtained and then processed to generate a non-redundant peptide list of antibody binding epitopes using the protein-based immunome wide association studies (PIWAS) algorithm. Each reconstructed 12-mer sequence was decomposed into 5- and 6-kmer components and log-enrichments of these shorter kmers were calculated at each amino acid position along a protein. We then determined epitope hits and seroreactive regions were based on the enrichment scores.

#### II. PCR amplification and sequencing

Plasmids were isolated for the selected bacteria and the genomic regions encoding the random peptides amplified with PCR. Sample-specific indexing primers were used to incorporate adaptors and indices for barcoding individual samples and to enable sample pooling for NGS sequencing and data demultiplexing after sequencing ^20,21^. Sequencing was done according to specifications of the Illumina NextSeq 500 using a High Output v2, 75 cycle kit (Illumina, FC-404-2005).

#### III. Data preprocessing using the PIWAS method

For each sample, approximately 750,000 to 3.6 million unique 12-mer sequences were obtained from the SERA assay and decomposed into constituent 5- and 6-mers ^12^. We used the published Protein-based Immunome Wide Association Study (PIWAS) ^22^ to detect enriched IgG signal in target pathogen proteomes (**Table S3**). IgG Enrichment scores for each *kmer* were calculated by dividing the number of unique 12-mers containing the *kmer* by the number of expected *kmer* reads for the sample, based on amino acid position composition of the sample. For each sample, the 5- and 6-mers were tiled against proteomes or protein sequences of interest. A tiling score was calculated at each amino acid (AA) on a protein based on the log-enrichment of the corresponding 5- or 6-mer enrichment scores, with a smoothing window size of five 5-mers and five 6-mers. That is, a PIWAS value at each AA position along a protein sequence represents an averaged score within a 5-AA frame using the tiling *z-*scores of 5-mers and 6-mers spanning the sequence. As a final step, we calculated normalized *kmer* enrichment values per AA position by normalizing the z-scores to a control population described in ^23^. The normalized z-scores are hereafter referred to as IgG enrichment values and represent antibody binding intensity. In *Supplementary Text* we show example alignments of individual-level normalized z-scores per AA position.

### Analysis of IgG enrichment values

#### I. Identification of linear protein-wide epitopes and seroreactive regions

Using the IgG enrichment values calculated above, an epitope in a protein was then defined as eight or more consecutive AA with a value above the 90^th^ percentile enrichment value for that protein (see **Fig 1** for illustration and **Supplementary Text** for real example data). We chose a conservative minimum length of eight as it has been suggested that linear stretches in epitopes are characteristically seven AA in length and antibody-binding energy is typically derived to about six residues ^24–26^. We then defined a seroreactive region, or an immunodominant region, as a contiguous protein fragment of high reactivity characterized by ≥5 overlapping epitopes at each AA position in that fragment. The relative start of a seroreactive region was the first AA position of the overlapping epitopes, after sorting the epitopes by their start AA residue. Accordingly, the relative end location was the last AA position of the overlapping epitopes sorted by length (see **Fig 1** for illustration and **Supplementary Text** for real example data). Therefore, the minimum length of a seroreactive region would be eight AA residues consistent with a stack of ≥5 overlapping epitopes. An occurrence of <5 overlapping epitopes at an amino acid position introduced a gap between seroreactive regions (**Supplementary Text**).

We limited protein-level summaries to seroreactive regions in order to focus inference on protein regions with maximum signal or enhanced reactivity, while ignoring infrequent epitope hits in the rest of the protein that could occur due to chance.

#### II. Definition of seropositivity, breadth and magnitude

A sample was seropositive to a protein, and by extension to a pathogen, if it had two or more epitope hits in the identified seroreactive regions. That is, after filtering for seroreactive regions only, seropositivity for a pathogen was defined as having at least two epitope hits across any of the queried proteins belonging to that pathogen. For example, a sample was seropositive for Rotavirus if it had two or more epitope hits in seroreactive regions of the VP4 protein alone, or collectively in seroreactive regions in VP4, VP7 and VP6 proteins. Assay specificity and sensitivity had been accounted for in the PIWAS analysis described above and therefore not needed to be considered for definition and interpretation of seropositivity.

The number of epitope hits was used as a proxy for breadth of immune response. For each serum sample, and for each evaluated pathogen protein, we defined breadth as *the number of epitopes located in seroreactive regions* and magnitude as *the average IgG signal intensity in the seroreactive regions*. We summarized breadth at each visit to assess population-level seroprevalence for various pathogens as described and shown in **Supplementary Text** and then narrowed our focus to six enteropathogens – (i) Norovirus, (ii) *Helicobacter pylori* (*H. pylori*), (iii) Rotavirus, (iv) adenovirus (HAdV), (v) heat-stable Enterotoxigenic *E. coli* (ETEC-ST) and (vi) *Salmonella enterica* serovar Typhi (typhoid, *S*. Typhi). Additionally, we included (vii) Measles virus and (viii) Poliovirus as positive controls to assess assay performance given that the age of the study cohort intersected with childhood immunization age.

### Statistical analyses

Statistical analyses were prespecified (https://osf.io/u92pc/). Children were analyzed according to their randomized group (intention-to-treat). The primary comparisons between the intervention and control groups focused on measurements in children ages 6 months or older, while accounting for repeated measures within each child. We excluded measurements at ages <6 months from the between-group comparisons because of the likely contributions of maternal IgG (prespecified). Secondary analyses compared treatment groups at ages 14 and 28 months separately to examine age-dependent differences in the intervention effect. For quantitative comparisons, we summarized data for each sample as – (i) count of epitopes in seroreactive regions per protein and cumulatively across all proteins for a pathogen, (ii) mean IgG enrichment value per protein calculated as sum of enrichment values divided by protein length.

Descriptive statistics were presented as frequencies and percentages; as median with interquartile ranges (IQRs); or mean with standard deviation (SD) or 95% confidence interval (CI). We estimated permutation *P-*values for differences between groups, permuting treatment at the cluster level (10,006 replicates). We report *P* values uncorrected and adjusted for multiple comparisons across pathogens using the Benjamini-Hochberg procedure with a 5% false discovery rate. All hypothesis tests were conducted with a two-sided significance level of *P* <0·05.

#### I. Intervention group comparison

Group comparisons combined information from ages 14 and 28 months. For each pathogen, we calculated differences in the number of epitope hits and mean IgG enrichment between treatment groups using a Wilcoxon rank sum test. We compared seroprevalence between groups by estimating prevalence difference (PD) using a linear-binomial regression model, and prevalence ratio (PR) using a generalized linear model framework with a log link and a Poisson distribution to improve convergence, plus robust standard errors (SEs) that treated individual study clusters as independent units.

#### II. Incident seroconversion

We assumed that all children were at risk of seroconversion at age 14 months on the basis that the period *between* visit 1 (median age, 3 months) and visit 2 (median age, 14 months) coincides with a period of declining maternal immunity. That is, if a child is seropositive at 14 months of age, it is unlikely to reflect maternal IgG. We only considered children who were seronegative at 14 months to be at risk of seroconversion at 28 months. Observations at 3 months of age (visit 1) were excluded in this analysis, and we did not anticipate seroreversion below age 27 months ^5^. Since seroconversion is an incident outcome, we estimated risk (cumulative incidence) and risk difference (RD) using a linear probability model with robust SEs at the cluster level. We estimated the ratio in seroconversion rate (or risk ratio, RR) using a log-binomial model with robust SEs at the cluster level.

### Mathematical models to reconstruct antibody dynamics

IgG measurements at an early age such as in this study, may include individuals with immunity from passive maternal transfer as well as induced by natural infection. These immunity sources are generally indistinguishable and can be separated under reasonable assumptions using catalytic models. For model fitting, we summarized the seropositives by age in months, resulting in discretized data with age range of 1 to 32 months. Given the relatively small number of samples and low seroprevalence for some pathogens, we consolidated seropositivity data from the intervention and control groups to increase the number of samples in each age class and allow for better binomial approximation of age-specific probability of infection.

For each pathogen, we fitted a modified catalytic model framework with an adaptable structure of maternal immunity to estimate (i) the force of infection defined as the rate at which susceptible individuals become infected (FOI, parameter λ), (ii) and the rate of losing maternal immunity (parameter γ). We assumed a constant λ over time and age, which estimates long-term averages of past force of infection.

We describe population dynamics of the model with the following closed system of differential equations as demonstrated in ^27^:

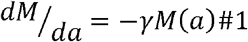

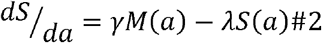

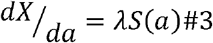

where *λ* > 0; *γ* > 0;

The system solution can be expressed as:

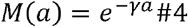

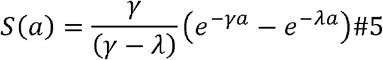

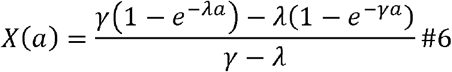

*M*(*a*) denotes the proportion of children of age a losing maternal immunity at a constant rate γ; *S*(*a*) denotes the proportion susceptible among children of age *a* born with maternal immunity; and *X*(*a*) denotes the proportion of children of age a that are seropositive from natural infection or vaccination.

For each age *a*, the accrued proportion of seropositive children and therefore the expected seroprevalence, *π*(*a*) = *M*(*a*) +*X*(*a*), can be calculated as 1 minus the corresponding susceptible or seronegative proportion:

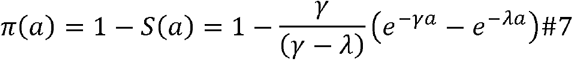

could also be interpreted as probability of being seropositive at
where 0 < *π* < 1.

The value *π* (*a*) could also be interpreted as probability of being seropositive at age a. The probability of infection per month unit time can be calculated as 1−e^−*λ*^. Population dynamics were reconstructed with the solutions listed above and using the posterior λ and γ mean estimates. We assumed all children are born with maternal immunity.

The seroprevalence data was modelled by binomial distribution with probability of success equal to *π*(*a*).

The models were implemented under a Bayesian model structure using cmdStanR (v.0.5.3). The choices of priors for λ and σ (**Table S4**), were based on prior predictive simulations of the age-profile of seropositivity, which showed a wide range of possible seropositivity profiles given our choice of priors (**Fig S2**). The priors for λ corresponded to an average force of infection between 0.5 to 0.9, while the half Cauchy distribution priors for γ allowed for a small decay rate of maternal immunity. The posterior distributions of the parameters were estimated using four chains, each with 3000 iterations and 900 burn-in. MCMC convergence was assessed by visual check of chain mixing and by the Gelman-Rubin R-hat diagnostic (threshold of <1.1). The effective sample size (ESS), the estimated number of independent samples accounting for autocorrelations generated by the MCMC run, was evaluated at a threshold of >400. Parameter estimates are presented as means with 95% credible intervals (CrIs) calculated from the 2.5% and 97.5 percentiles of the posterior distributions.

## RESULTS

### Group analysis of WASH and nutrition intervention effect on immune response

We analyzed sera from 120 children randomized to the intervention (WASH+N) and control groups, 60 children in each group. There was no significant difference in the distribution of epitope hits between the intervention and control groups (Wilcoxon rank sum test, *P* >0.05), and in the mean seroprevalence (**Table 1**). Across pathogens, the mean IgG enrichment levels were lower in the intervention group versus the control group but were not significantly different between groups after adjustment for multiple comparisons (**Table S5**). Since antibody responses are considered a measure of cumulative exposure over time, seroprevalence estimates at the median ages of 14 and 28 months were interpreted as a cumulative incidence. The overall seroconversion was lowest in *S*. Typhi (8%, 4%-11%), and highest in Norovirus (98%, 96%-100%), heat-stable producing enterotoxigenic *E. coli* (ETEC-ST) (98%, 96%-100%) and adenovirus (100%) (**Table S6**). The difference in risk or cumulative incidence and the risk ratio between groups was not significant among the pathogens assessed (*P* >0.05) (**Table S7**). For ETEC-ST, the children in the intervention group had a lower relative risk of seroconversion than children in the control group (risk difference: 3%) though not significant (**Table S7**).

**Table 1.**
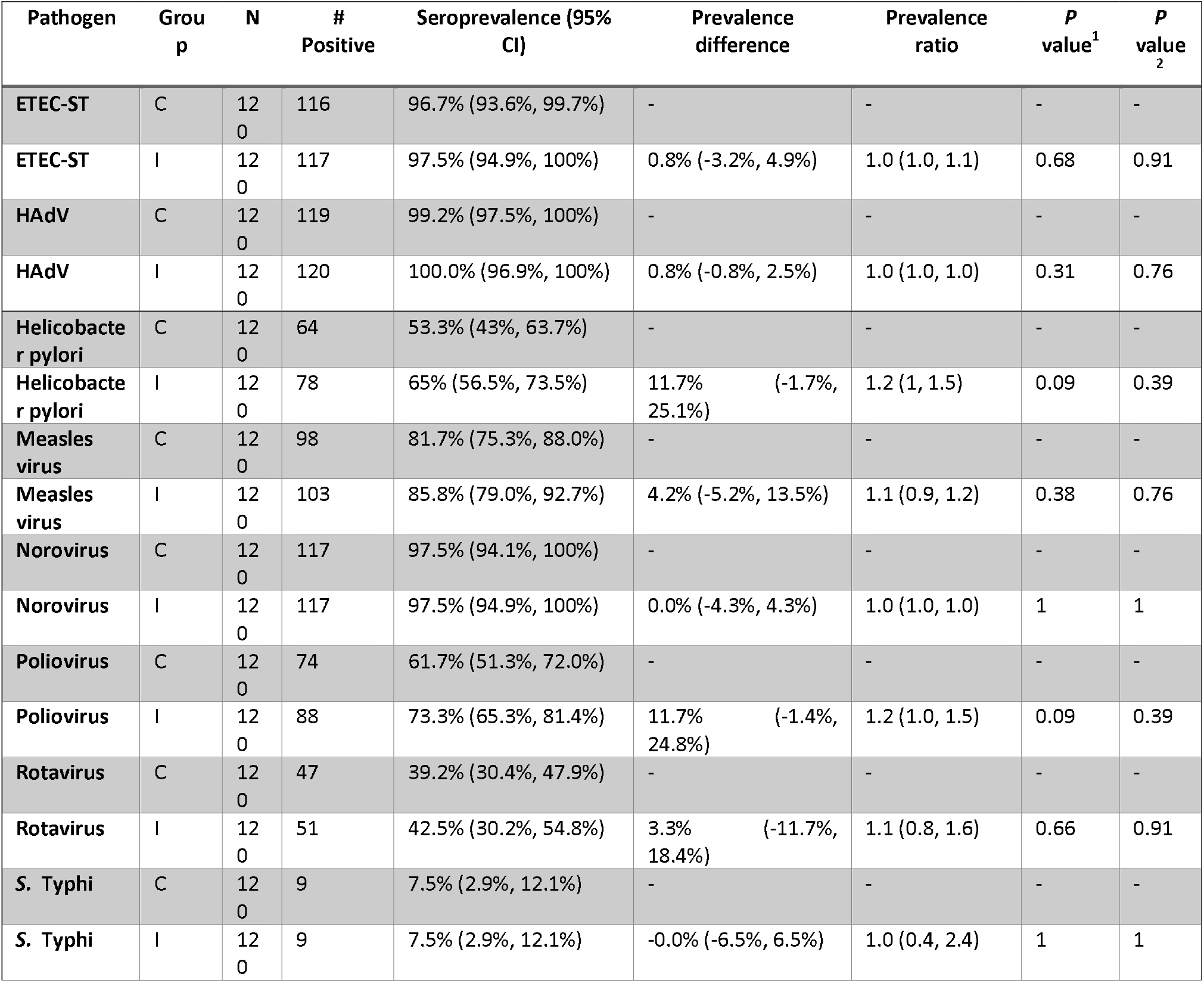
Estimates are based on 60 children in each group measured twice at median ages 14 and 28 months. 95% confidence intervals (CIs) and *P*-values^1^ account for repeated measures within children and clusters using robust standard errors. *P*-values^2^ are the Benjamini-Hochberg FDR-adjusted p-values for multiple comparisons. The control group was the reference in the regression analysis. Key: C = control, I = intervention.

### Magnitude and breadth of antibody response

Collectively, we analyzed antibody reactivity in 360 serum samples (60 children x 3 visits per child x 2 groups) across 118 proteins in 8 pathogens, ranging from a single protein to 34 proteins per pathogen (**Table S3)**. For eight example proteins, one for each pathogen listed above, the average IgG signal was similar among sampling visits but slightly higher in the intervention group (**Fig 2, Table S5**), and along the protein length (**Fig S3 and Fig S4)**. Yet, there were varying levels of IgG signal enrichment across pathogen proteomes and in some protein regions the antibody reactivity changed markedly between the sampling timepoints (**Fig 3)**. Waning of maternal immunity was seen as a drop in the mean IgG enrichment value – the green line below zero in **Fig 3**, while a boost in immune response due to infection or vaccination was seen as increase in the mean IgG enrichment value between 14 and 28 months – the red line in **Fig 3**. Rotavirus VP7 and *H. pylori* CagA proteins are good examples. We used paired *t*.*test* and multiple comparison adjustment methods to determine whether the change in mean IgG enrichment values was significantly different from zero. In the *S*. Typhi HlyE protein, IgG reactivity from age 3 to 14 months was notably enhanced in the first 100 AA (adjusted *P* <0.05) and then dampened in the remaining protein fragment (**Fig 3**). The magnitude of IgG signal per sample increased with cumulative breadth or immune repertoire diversity measured as the total number of unique epitope hits per sample (**Fig 4**). This was more prominent for pathogens with a larger number of protein components evaluated, e.g., ETEC-ST, HAdV, Measles virus and Norovirus, which are therefore more likely to appear to have a more diverse antibody repertoire.

**Fig 2.**
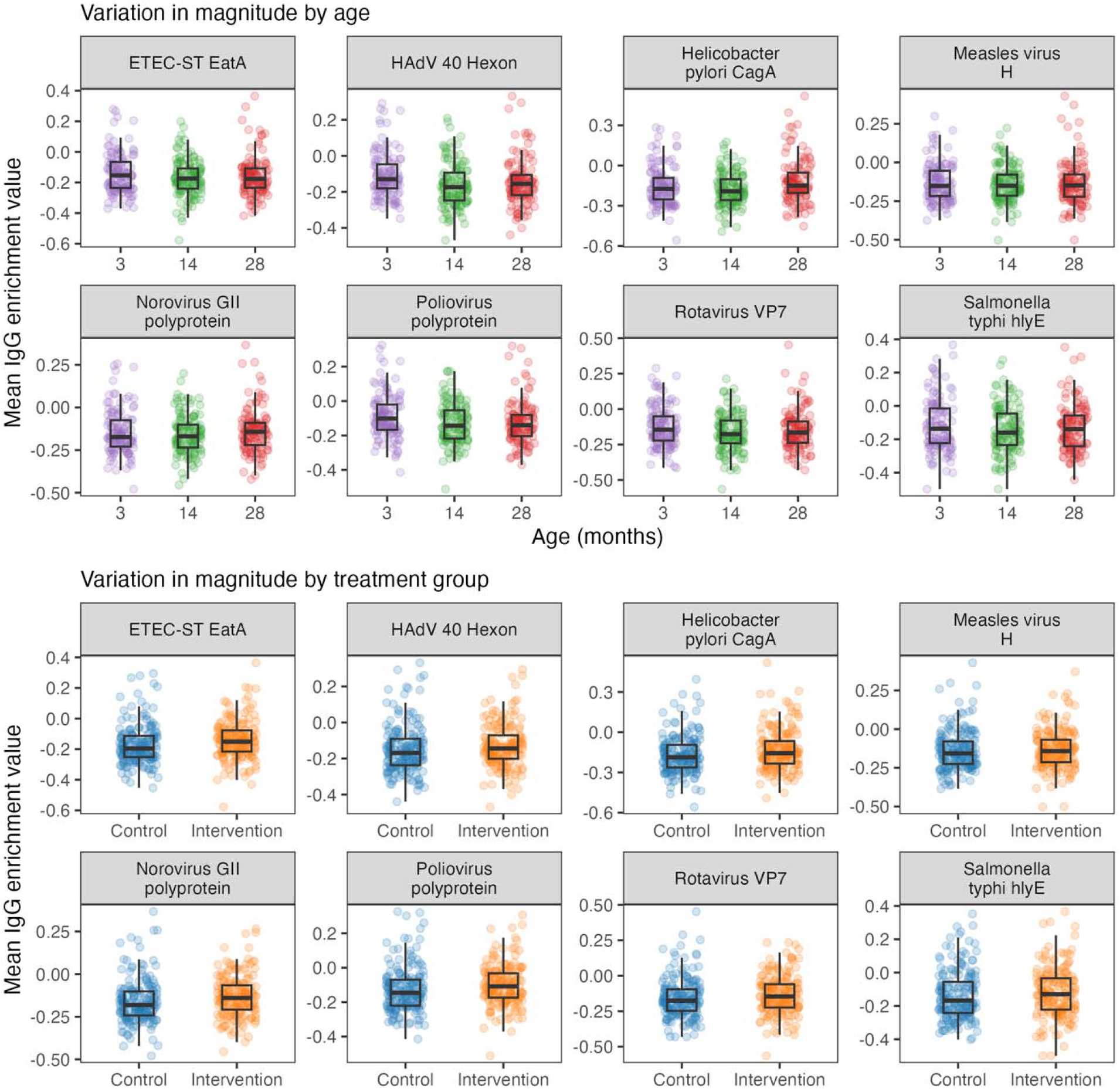
Averaged IgG enrichment scores at individual level. For each sample, the IgG enrichment scores were summarized as means by calculating the sum of enrichment values across the protein length and dividing by protein length (number of amino acids per protein). The data summaries are stratified by sampling time and treatment group assigned during recruitment. Here we show data for eight example pathogen-protein pairs.

**Fig 3.**
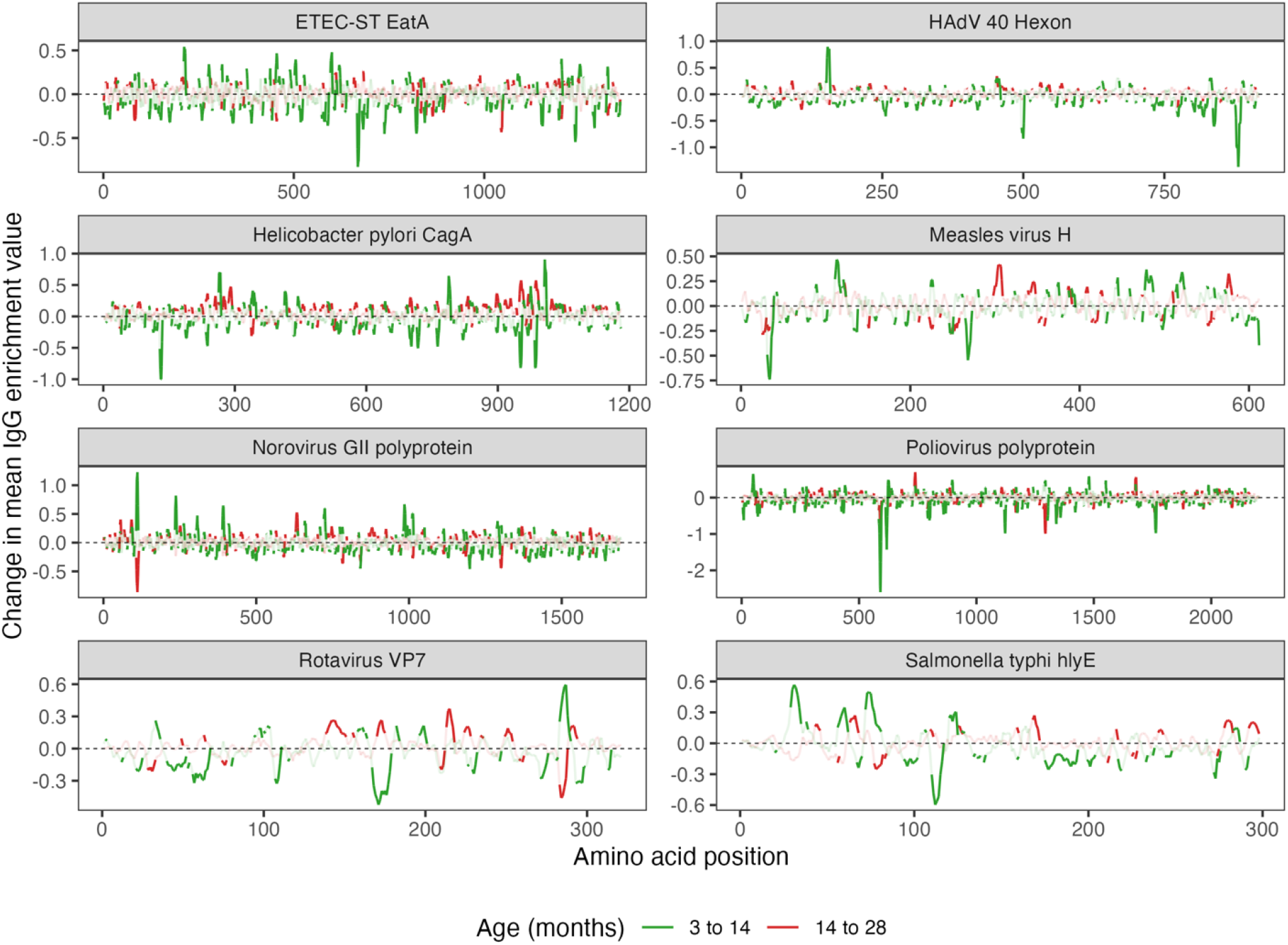
Averaged changes in IgG enrichment over time. For each amino acid (AA) position we calculated the average enrichment value at median ages 3, 14 and 28 months and then subtracted those average values to determine the change in IgG enrichment over time. That is, [*IgG*_14_ - *IgG*_3_] in green and [*IgG*_28_ – *IgG*_14_] in red. Values below 0 (the black dashed line) indicate a decrease in enrichment and values above 0 indicate an increase in enrichment. We used paired *t*.*test* and multiple comparison adjustment methods to determine whether the changes were significantly different from zero. A significant change (Bonferroni-Hochberg adjusted *P* <0.05) is shown as a full colored line and while the faded color indicates non-significant change (adjusted *P* >0.05).

**Fig 4.**
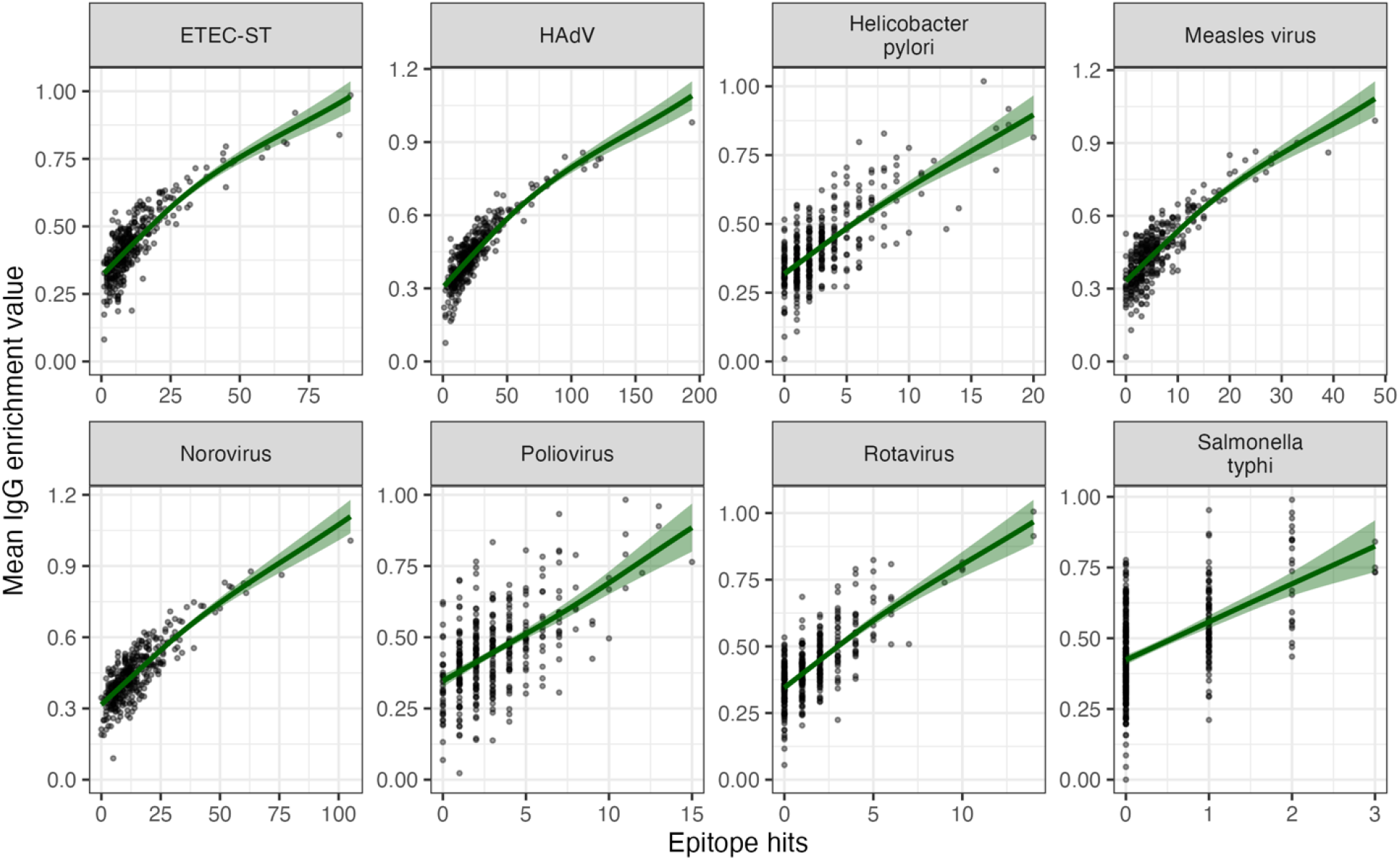
Averaged IgG enrichment versus antibody breadth. The figure illustrates association between individual-level antibody reactivity versus breadth (total count of epitope hits in a protein). For each sample, the mean IgG enrichment score was calculated across the entire protein length – each panel has 360 datapoints (= 60 children per group x 2 groups x 3 sampling visits). The dark green lines show a loess smoothing function, and the shading shows 95% confidence intervals.

Generally, there was minimal age effect in breadth (number of epitope hits) and magnitude (mean IgG enrichment value) (**Fig S5** and **Fig S6**). In the eight pathogens evaluated, there were no significant changes (adjusted *P* value >0.05) in breadth and magnitude across age except for Poliovirus. At the individual level, serum reactivity and specificity were diverse across pathogen as shown in **Fig S7** and **Fig S8**, where AA residues and protein regions with enhanced seroreactivity are captured as peaks in the IgG enrichment value and by multiple overlapping linear epitope hits, respectively. We observed that the inferred seroreactive regions spanned large protein fragments or nearly full protein length (**Fig S8**). For each pathogen, we summarized breadth as crude seroprevalence by calculating the proportion of seropositive children at each sampling visit. Overall, seroprevalence was similar over age or time and uniform (did not differ significantly) between treatment groups (**Fig S9**).

We also looked at the antibody reactivity of Norovirus, ETEC-ST, Measles virus and Poliovirus in slightly more detail. The Norovirus major (VP1) and minor (VP2) capsid proteins were broadly reactive along the protein length in both genotypes (**Fig S10** and **Fig S11)**, which suggests antigen homology resulting in highly correlated antibody responses, though we observe a marginally attenuated seroreactivity in Norovirus genotype II compared to genotype I. For genotype I, the more enhanced antibody reactivity occurred near the C-terminus in both capsid proteins and there was more seroreactivity at the central and C-terminus for VP1 and at the N-terminus for VP2 for genotype II. There was high serum reactivity to ETEC-ST plasmid-encoded antigens that are involved in intestinal colonization, characterized by large clusters of epitopes and seroreactive regions (**Fig S12** and **Fig S13**), though it did not seem to vary by intervention group or age in the six ETEC-ST antigens.

Immune responses to vaccine-targeted pathogens were considered as a positive control – vaccinated children should have antibodies to these pathogens. For both Measles virus and Poliovirus, there was no difference in the magnitude and breadth of response between groups, but there was higher seropositivity for the Measles virus attachment glycoprotein hemagglutinin (H) compared to the fusion glycoprotein (F) (**Fig S14**). The intervention group was more seropositive for the Measles virus F glycoprotein, while the control group was more seropositive for H protein (**Fig S14**). The overall cumulative incidence of seroconversion for Measles virus and Poliovirus was estimated at 82% and 67%, respectively (**Table S6**), and the population-level model estimates showed a peak in susceptibility at three to six months of age, as discussed below.

### Model estimates of age-dependent immunity dynamics

We implemented a modified catalytic model framework that incorporates maternal immunity to infer antibody dynamics and the expected population immune profiles. We observe substantial variation in the inferred antibody dynamics across pathogens. The inferred cumulative proportion seropositive, *π*(*a*) = *M*(*a*) +*X*(*a*) (**Methods**), showed the expected profile – that is, a decline then steady rise with increasing age – which was most evident for *H. pylori*, Measles virus, Poliovirus and Rotavirus (blue lines, **Fig 5**). ETEC-ST, HAdV and Norovirus had a very high estimated cumulative seropositivity, with nearly 100% seropositivity across all study ages (1-36 months). We estimated a slower rate of maternal antibody decay for ETEC-ST, HAdV and Norovirus, which had ≥60% seroprevalence at six months of age, and 40% seroprevalence at 12 months. For Measles virus and Rotavirus, the maternal antibody levels at the age of six months were estimated at 50% and 18%, respectively. At 12 months, seroprevalence due to infection and/or vaccination was estimated at 20% for ETEC-ST, Norovirus, Rotavirus and H. pylori; and at 30% for HAdV and Measles virus (**Fig 5**). For *S*. Typhi, the immunity profiles were non-differentiable and the overall *S*. Typhi seroprevalence remained well below 20% in the study cohort and only very slowly increased by age, suggesting limited exposure in this age range for this pathogen.

**Fig 5.**
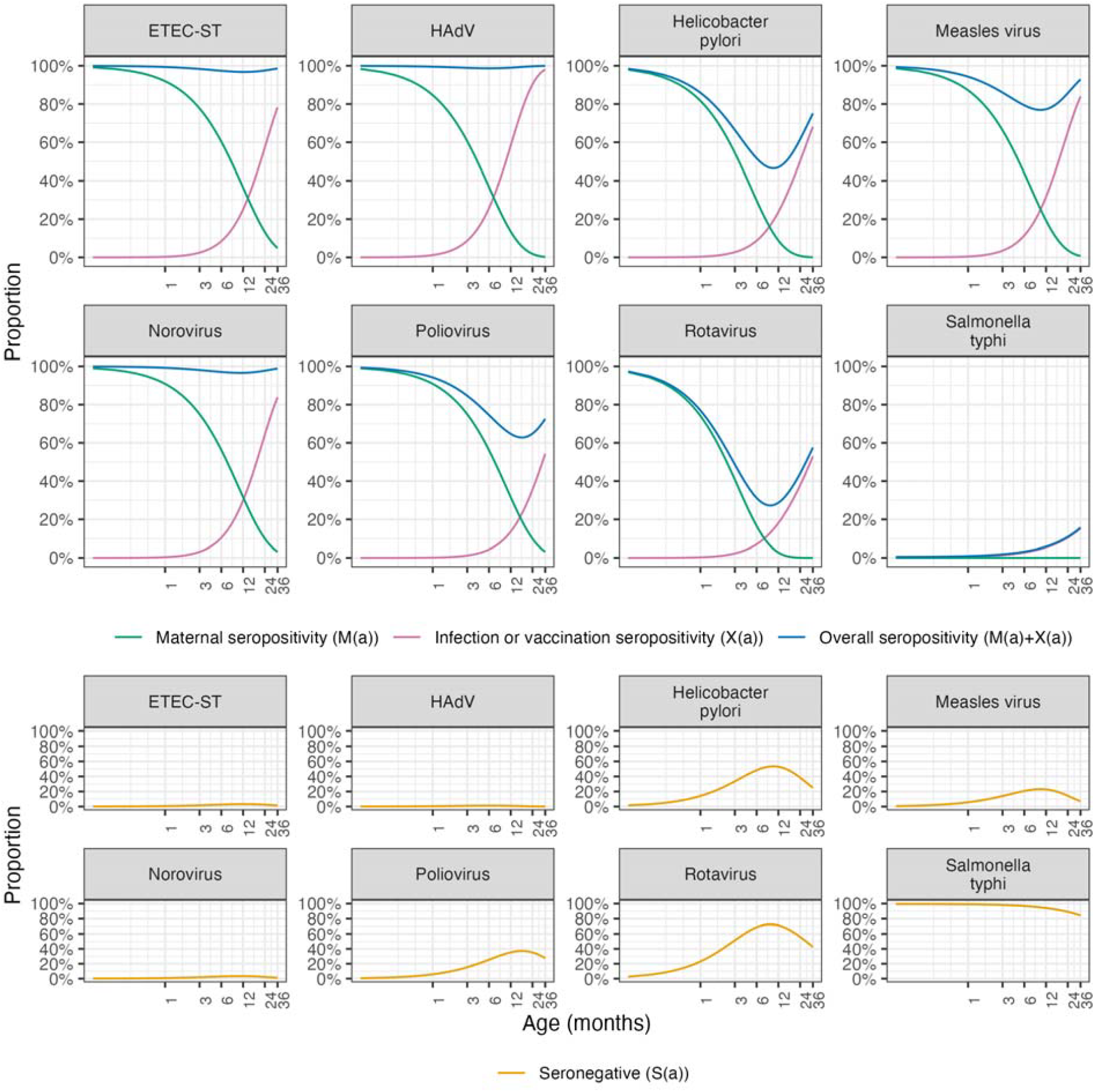
Simulated expected population immune profiles. The figure illustrates the predicted population age seroprevalence profiles based on model parameter estimates. The color of the line denotes the immunity profile: <u>green</u> line represents the inferred proportion with maternal antibodies; <u>pink</u> represents the inferred proportion seropositive from infection or immunization (Measles virus and Poliovirus); <u>blue</u> indicates cumulative seropositivity and <u>orange</u> (bottom panels) represents the proportion seronegative. Note that the age axis is log-transformed for better visualization in the much younger age classes.

We estimated variation in the mean duration of seropositivity due to the presence of maternal antibodies (1/γ) across pathogens, with the longest duration being 12 months (95% Credible Interval [CI], 7-24) for ETEC-ST, followed by Norovirus (10 months (95% CI, 6-21)) and Poliovirus (10 months (95% CI, 7-15)). The durations of seropositivity due to maternal antibodies for Measles virus and Rotavirus were estimated at 7 months (95% CI, 5-11) and 3 months (95% CI, 2-4), respectively. For *S*. Typhi, the model estimated a mean duration of maternal seropositivity of <1 month, which could be attributed to the very low overall seropositivity – the *S*. Typhi data were not sufficient to estimate both the force of infection and maternal antibody decay. In all cases, the model dynamics assumed all births have maternal antibodies, that is, *M* (*a* = 0) = 1. This is a crude assumption and might not be applicable to all pathogens.

At 36 months of age, we estimated cumulative seropositivity of 95% for Measles virus, 75% for *H. pylori* and Poliovirus, and 60% for Rotavirus. The proportion of susceptible or seronegative children, a), was calculated as the complement of the cumulative proportion seropositive, *S*(*a*) =1 − *π* (*a*). We estimated 50% to 80% susceptibility to Rotavirus in the age interval of 3-24 months, and above 80% for *S*. Typhi across the entire age range of 1-36 months (orange lines, **Fig 5**). All parameter estimates are presented in **Table S8**. The model exhibits a good fit to the observed data across all eight pathogens and captured the underlying pattern of decline then subsequent increase in seroprevalence (**Fig 6**). The estimated seroprevalence closely matches the observed data except for the very young (ages 1 and 2 months) for Poliovirus and Rotavirus.

**Fig 6.**
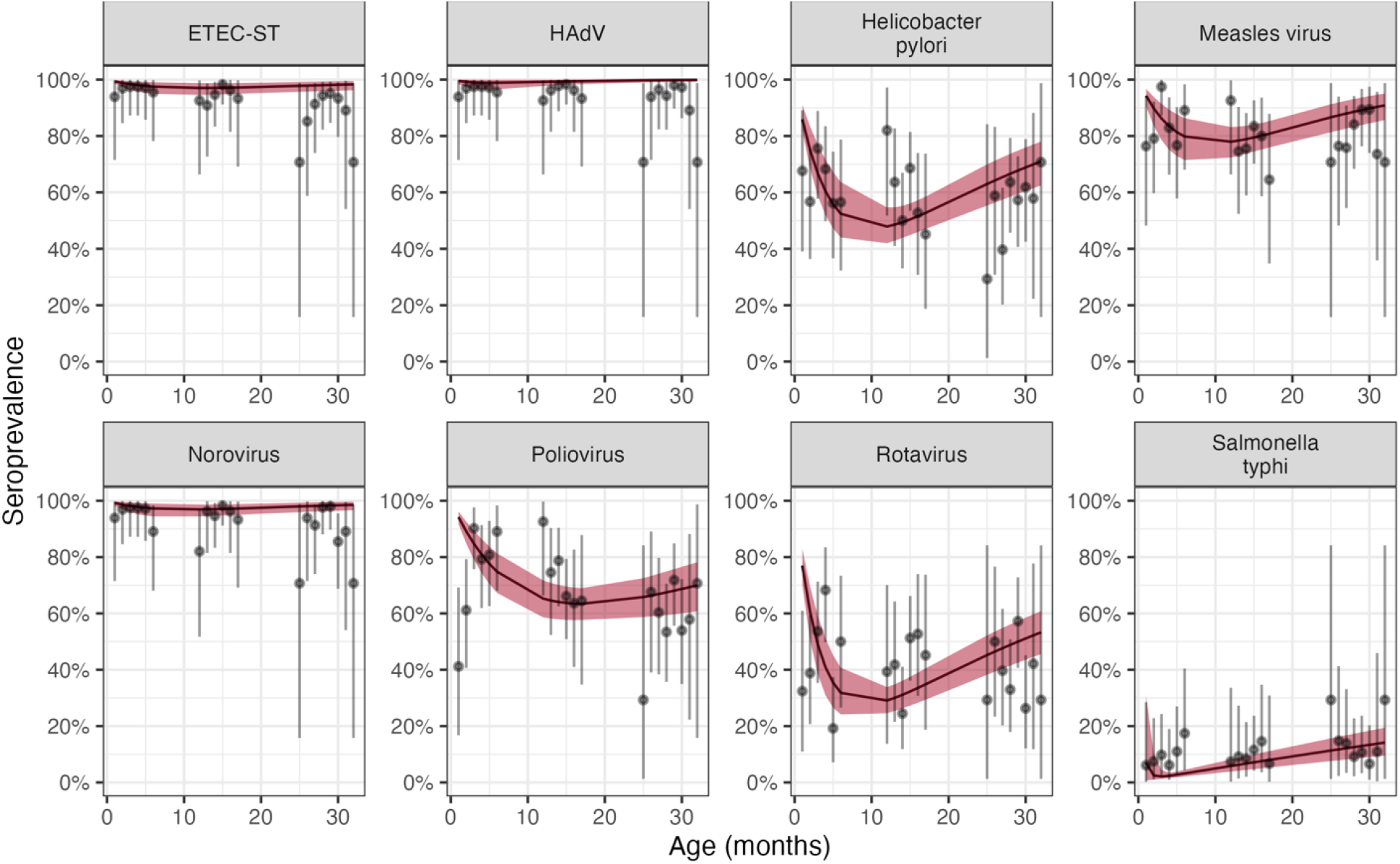
Model fitting to data. Observed (gray) and estimated (red) seroprevalence by age. The gray points and intervals indicate the point estimate and 95% binomial confidence intervals around the observed proportion of seropositivity. The colored lines and ribbons indicate the mean and 95% credible intervals of the posterior estimates of seroprevalence.

## DISCUSSION

We used a highly-multiplexed peptide library screening to characterize humoral responses to diverse pathogen types and species. The method yielded high resolution antibody binding data at amino acid level that we used to identify putative antigenic epitopes and regions of elevated seroreactivity. Overall, we observed broad individual and population level antibody reactivity to enteric pathogen proteomes characterized by varied signal intensity across the protein length as well as localized protein fragments of enriched reactivity from which we identified putative epitope variants. We determined conserved IgG binding activity across age – that is, antigenic epitopes and regions of amplified seroreactivity which remained consistent at the three sampling timepoints.

WASH+N intervention and control groups had similar patterns of seroreactivity and there were no notable or significant differences between groups in terms of IgG breadth or magnitude of response, which either indicates a real absence of immunological signal in the sampled age group or insufficiency of the method used. A previous analysis of PCR detected infections in stool from the same cohort detected benefits of WASH and nutrition improvements on enteric virus and protozoan infections ^19,28^.

We had hypothesized that the intervention group would have less exposure or experience fewer infections and therefore demonstrate lower antibody reactivity, with IgG providing a more integrated measure of cumulative exposure compared with a single stool sample. There may have been an effect of WASH on interrupting pathogen transmission as noted both in reported diarrhea and stool pathogen assessment, but the bacterial display screening method was unable to detect these differences sufficiently via serology and across age. Other analyses of this cohort suggested that WASH+N intervention enhanced immunoprotective and immunoregulatory responses, and suppressed or counteracted the inflammatory and immunopathological responses ^29^.

Antibodies to Rotavirus VP7 surface glycoprotein protect against virus infectivity ^30^ and were strongly enriched in the WASH Bangladesh samples. It is important to note that these samples preceded introduction of Rotavirus vaccine in Bangladesh, so IgG reflects natural infections (not vaccination). We determined potential immunodominant epitopes mostly in the central VP7 protein region, around amino acids 50 to 200 (**Fig S8**). This region contains antigenic site A composed of amino acids 87 to 96, which may be involved in the formation of a major protective epitope and cross-reactive neutralization ^31,32^. Our samples were enriched for anti-Measles virus antibodies that recognized known immunodominant linear epitopes in the H protein, including amino acid residues 244-250, 188-199 and 190-200. The epitope at 244-250 is designated as ‘the neutralizing epitope’ and residues 188-199 comprise the ‘receptor binding epitope’ ^33^. All three epitopes are targets for neutralizing Measles virus infection ^33^.

Norovirus capsid proteins VP1 and VP2 are crucial antigenic components, likely to modulate immune responses ^34^. Serum samples were highly seroreactive to VP1 and VP2 as depicted by numerous predicted overlapping linear epitopes (**Fig S10**). Norovirus antibody recognition remained high between visits 1 (median age, 3 months) and 3 (median age, 28 months), suggesting persistent boosting as a function of frequent exposure. This aligns with a recent study in Nicaragua that reported new infections in early infancy, with children between 3 and 12 months of age experiencing ≥4-fold increases in Norovirus IgG antibodies ^35^. We estimated waning of Norovirus maternal antibodies reaching nearly 50% seroprevalence by six months of age, which is also consistent with seroprevalence in the Nicaraguan birth cohort ^35^.

We estimated a low seroprevalence for *S*. Typhi HlyE protein (**Fig 5 and 6**), which could imply fewer infections and high susceptibility in the age group in this study. We estimated an overall high seroprevalence for the enterotoxigenic *E. coli* (ETEC-ST) (**Fig 5 and 6**), and even though a previous study suggested high sequence homology between *E. coli* HlyE and *S*. Typhi HlyE ^36^, we do not expect reciprocal cross-reaction between the two bacterial pathogens based on these antigens. From the IgG enrichment data, we predicted a linear epitope between amino acids 175 to 186, which overlaps with a known *S*. Typhi HlyE B-cell epitope at amino acids 180 to 188 ^36^. We also predicted several other linear epitopes containing amino acid residues PYSQESVLSADSQNQK that constitute a known conformational B-cell epitope. These B-cell epitopes were shown in a published to have higher IgA and IgG reactivity, compared to IgM, with potential role in intestinal mucosa immune response ^36^.

Antibody prevalence in early childhood, as in our study cohort, could be a combination of maternal antibody persistence and presence of antibodies acquired due to direct pathogen exposure. We applied Bayesian catalytic models to estimate age-specific seroprevalence considering the influence of maternal antibodies. As expected, maternally derived immune dynamics decline exponentially from birth as a function of age, consistent with the recognized underlying mechanism ^37^. Our findings suggest similar age patterns across pathogens, where seroprevalence decreases between zero and four months and then increases steadily from six months reaching 100% by 36 months of age (**Fig 5**). For all pathogens except *S*. Typhi, we estimated antibody acquisition to a higher level than the presumed maternal immunity (the seroprevalence in the first month), which suggests that by age three years children will have attained antibody prevalence proportional to the level circulating in adults. This is not far-fetched and has been observed for enteroviruses and Norovirus ^35,38–40^. The low prevalence and short duration of maternal antibodies estimated for *S*. Typhi is unlikely for adults living in an endemic setting, although plausible for a rural setting like where the trial was conducted ^41^. Even so, it is possible there was still high *S*. Typhi transmission, but the SERA assay was inadequate to capture the conformational immunodominant epitopes of HlyE.

We estimated a median duration of maternal immunity of 10 months (range, 1-11 months), shortest for *S*. Typhi and longest for Norovirus and Rotavirus. While we did not quantify antibody titers using antigen-specific assays such as ELISA or multiplex bead assays, our inferred overall trends in antibody decay suggest risk of infection in the first year of life and are consistent with the existing knowledge of duration of protection in infants and young children ^37^. Still, lack of detection of peptide enrichment for several pathogens e.g., intestinal protozoa, *Campylobacter jejuni* and *S*. Typhi, should not be concluded as absence of the corresponding antibody specificity.

Our study had limitations. The samples were not evenly distributed by age resulting in small numbers at some specific ages, which could influence the precision of age-specific seroprevalence estimates. For instance, only eight children were 12 months old, only one was 25 months old, and just five were 31 months old. The primary comparisons between intervention groups were aggregated over 120 samples in each group providing reasonably good statistical power, but we refrained from estimating age-specific seroprevalence by intervention group. We also did not explore how factors such as birthweight, maternal education, nutrition or maternal comorbidities might have affected serum reactivity, and this could be a topic for additional study. Nonetheless, our study demonstrates the use of high-throughput antibody screening to characterize age-specific antibody profiles and provides a motivation for further epitope-level serological analysis of the humoral response to enteric infections. Further studies are warranted to confirm the breadth of immune response and investigate protection from infection or illness.

In summary, we show the potential of random sequence peptide arrays probed with human sera containing antibody mixture of unknown complexity to measure endpoints in an RCT, e.g., children’s exposure to enteric pathogens in the household environment. In future RCTs, using protein array screening with more frequent sampling or larger cohorts might enable the detection of subtle differences or dynamic changes between clinically well-characterized intervention and control groups. Peptide display systems are still being developed and improved, and new immunological and biomedical applications can be expected to become available for disease surveillance and evaluating trial outcomes.

Our bioinformatic approach for identification of antibody binding epitopes is particularly notable as we identified antigenic peptides reported in other studies. Our results demonstrate further the potential of proteome-scale antigen analyses for multiplex testing of a wide array of pathogens of epidemiological and clinical interest, and for robust estimation and resolution of humoral response not readily achieved with traditional IgG assays.

## Acknowledgements

The authors would like to acknowledge Dr. Christine Tedijanto for input on the analysis plan.

## Funding

National Institute for Allergy and Infectious Diseases grant R01AI166671 (BFA).

## Author contributions

Following CRediT taxonomy, conceptualization (RCC,DTL,AL,BFA), data curation (EK,NW,MZ,KK,JR,JS), formal analysis (EK,NW,BFA), funding acquisition (BFA), investigation (SA,MdZR,AKS,SLF,SA,MdSH,PM,MR,ANM,SL,AL,BFA), methodology (EK,NW,MZ,KK,JR,JS,BFA), project administration (BFA), resources (SL,AL), software (EK,NW,BFA), supervision (BFA), validation (EK,NW), visualization (EK,BFA), Writing – Original Draft Preparation (EK,BFA), Writing – Review & Editing (all authors).

## Competing interests

Authors declare that they have no competing interests.

## Data and code availability

All data needed to evaluate the conclusions in the paper are present in the paper and/or the Supplementary Materials. All data displayed in the figures and used for analyses are available at (https://osf.io/u92pc/). All code used for analysis are available at (https://osf.io/u92pc/).

## Notes

### Competing Interest Statement

The authors have declared no competing interest.

### Clinical Trial

NCT01590095

